# Machine Learning Prediction of Autism ADOS-2 Calibrated Severity Scores Using Genetic and Behavioral Features

**DOI:** 10.64898/2026.11.20.26368220

**Authors:** Nithin Aditya Pradeepkumar, LeeAnne Green Snyder, Brett Trost, Bridget A. Fernandez, Chang April Shu

## Abstract

**Background:** Autism spectrum disorder (ASD) is a heterogeneous neurodevelopmental condition characterized by social communication difficulties and restricted, repetitive behaviors. While machine learning has been widely applied in autism research, few studies have predicted continuous autism severity measures such as the Autism Diagnostic Observation Schedule Calibrated Severity Score (ADOS CSS). We evaluated behavioral and genetic predictors of ADOS CSS across ADOS modules.

**Methods:** Participants from the Simons Simplex Collection who completed ADOS Modules 1, 2, or 3 were included. Behavioral questionnaire data and gene-level measures of loss-of-function intolerance (pLI, LOEUF) and brain-expression specificity (pExt) were used to train 12 machine learning models separately for each module. Performance was evaluated using mean absolute error (MAE), Pearson correlation (r), and SHAP analyses.

**Results:** Ensemble-based models achieved the strongest performance across modules. In Module 1, adding genetic predictors improved Random Forest performance (MAE 1.189 to 1.051; r 0.204 to 0.340). Module 2 achieved the highest overall performance, with XGBoost reaching r ≈ 0.49 and MAE = 1.010. SHAP analyses showed that genetic predictors contributed most strongly in Module 1, whereas predictions for Module 3 were driven primarily by verbal, behavioral, and social-cognitive features.

**Conclusions:** Integrating genetic and behavioral features improved ADOS CSS prediction, particularly in children with limited expressive language. Genetic information was most informative in Module 1, while behavioral and social-cognitive measures became increasingly important in later developmental stages, consistent with evidence that genetic burden is more strongly associated with greater developmental impairment.

## 1. INTRODUCTION

Autism Spectrum Disorder (ASD) is a complex neurodevelopmental condition that affects how individuals communicate, interact with others, and engage in repetitive behaviors. Autism symptoms vary greatly in both type and severity. This variability has motivated the development of standardized tools that can capture autism symptom severity across the spectrum. One of the most widely used such tools is the Autism Diagnostic Observation Schedule (ADOS-2) (Lord et al., 2012).

The ADOS involves structured tasks carried out with scoring systems that capture behaviors relevant to autism. To allow comparison across individuals of different ages and language abilities, the authors (Gotham et al., 2009) developed the ADOS Calibrated Severity Score (CSS) or comparison score. The CSS places scores on a standardized 1–10 scale that reflects autism symptom severity. It is useful for both clinical and research purposes. We use the term “severity” throughout this manuscript. This is consistent with the terminology established in the original ADOS CSS literature. Our use of this term solely reflects the standardized instrument and its source literature.

Obtaining ADOS scores at scale can be a challenge in large population studies, because ADOS assessments require in-person evaluations by trained clinicians. For this reason, many large datasets rely on parent or caregiver-reported behavioral questionnaires that can be administered efficiently and at scale, such as the Social Responsiveness Scale (SRS; Constantino et al., 2005) and the Repetitive Behavior Scale–Revised (RBS-R; Bodfish et al., 2000).

Genetic factors are known to play a major role in ASD. Twin and family studies have consistently shown high heritability estimates, ranging from 64% to 91% (Tick et al., 2016). Large-scale genetic studies have identified common variants associated with ASD (Grove et al., 2019). Other work has shown rare variants and copy number variants (CNVs) that are often linked to more severe forms of the condition (Trost et al., 2022; Zhou et al., 2022). This evidence suggests that incorporating genetic data may offer prognostic information about the potential trajectory of ASD for that person. This differs from an ADOS assessment alone, which involves several hours of direct clinical assessment on a single day.

Machine learning (ML) prediction of autism diagnosis and severity has gained interest in the field, supported by both methodological advances in prediction accuracy and its value in assisting with diagnostic validation in large-scale research studies. Early studies demonstrated the feasibility of ML for reducing questionnaire item sets and detecting ASD or differentiating it from related conditions with overlapping features such as ADHD (Duda et al., 2017). This illustrates the potential of data-driven feature selection. Recent work has expanded the scope of ML applications for autism detection. Farooq et al. (Farooq et al., 2023) applied federated learning to ASD classification, training logistic regression and support vector machine models on distributed datasets of children and adults, and demonstrated high accuracy for early detection. More recently, Rajagopalan et al. (Rajagopalan et al., 2024) developed an ML framework (AutMedAI) using minimal medical and background information from over 30,000 participants in the SPARK cohort. Their gradient boosting model achieved strong predictive performance (AUROC ≈ 0.90) and generalized well to external datasets, including the Simons Simplex Collection. Briguglio et al. (2023) used maternal and infant clinical history, including gastrointestinal symptoms, EEG findings, and sleep disturbances, to predict ADOS-2 domain scores and distinguish ASD from multi-systemic developmental disorder in young children. Together, these studies show the potential of ML approaches for scalable ASD detection, whether using advanced frameworks such as federated learning or minimal-feature classifiers on population-level data.

Despite this progress, very few studies have attempted to predict standardized measures of autism severity such as ADOS CSS using other sources of information. Most prior work has focused either on binary classification (ASD or non-ASD) using feature reduction, or on identifying latent phenotypic subgroups, leaving a gap in approaches that can provide continuous, comparable measures of ASD core symptom severity. The goal of this study is to develop a machine learning pipeline that predicts ADOS CSS using both behavioral questionnaires and genetic features. We used a clinically and genetically well-characterized ASD cohort, the Simons Simplex Collection (SSC), to develop machine learning models to predict ADOS CSS for modules 1-3. We compared multiple machine learning models trained on behavioral questionnaires alone, and behavioral questionnaires combined with genetic data, and assessed how genetic data improves the prediction performance. We also determined which behavioral and genetic features contribute the most to predicting ADOS CSS. To our knowledge this is the first study to examine how integrating behavioral questionnaires with genetic features influences the prediction of ADOS CSS.

## 2. METHODS

### 2.1 Study samples

Data was obtained from the Simons Simplex Collection (SSC) (Fischbach & Lord, 2010), a core project of the Simons Foundation Autism Research Initiative (SFARI). The SSC contains phenotypic data from more than 2,600 simplex families, defined as those with exactly one child with ASD, as well as his or her unaffected parents and siblings.

Standardized instruments included: the SRS, which measures social communication and behavioral impairment (Constantino et al., 2005); the Repetitive Behavior Scale-Revised (RBS-R), which assesses restricted and repetitive behaviors (Bodfish et al., 2000). the Vineland Adaptive Behavior Scales - Parent/Caregiver Interview form, which evaluate adaptive functioning across communication, daily living skills, and socialization (Sparrow et al., 2005). and the Aberrant Behaviour Checklist (ABC; Aman & Singh REF), which captures behavior problems and repetitive features often associated with autism and developmental disabilities.

The Calibrated Severity Score (CSS) is a standardized metric derived from the Autism Diagnostic Observation Schedule (ADOS). It quantifies the core symptom severity of autism in a way that is consistent across different age and language levels. It was originally proposed by Gotham et al. (2009). The CSS was developed to address limitations of the ADOS raw total score. Raw totals vary across modules. They are also heavily influenced by factors such as verbal IQ, age, and language development. The CSS maps raw ADOS totals onto a 10-point standardized scale. Specifically, it maps the sum of the Social Affect (SA) and Restricted and Repetitive Behaviors (RRB) domain scores. Calibrated Severity Scores are divided into four broad descriptive levels. Scores of 1 to 2 indicate minimal symptoms. Scores of 3 to 4 indicate low severity. Scores of 5 to 7 indicate moderate severity. Scores of 8 to 10 indicate high severity. This calibration enables direct comparison of the severity of core autism features across modules and over time. It is especially valuable for longitudinal and cross-sectional research. Empirical studies, including Shumway et al. (2012), have shown that CSS values are more uniformly distributed across diagnostic categories than raw ADOS scores. CSS values are also less dependent on verbal ability. They demonstrate good temporal stability over 12 to 24 months. Consequently, the CSS serves as a more reliable and valid index of autism symptom severity for research and clinical applications.

### 2.2 Genetic Data

Genetic information for the SSC cohort was obtained from large-scale sequencing efforts. These efforts identified *de novo* variants across the genome. Specifically, we relied on *de novo* variant data reported by Zhou et al. (2022), which analyzed 42,607 autism cases, including SSC participants. Their analysis was performed on exonic and coding regions of the genome.

Each participant in our sample carried a single qualifying *de novo* variant. This variant was either a single-nucleotide variant (SNV) or an insertion/deletion (indel). No population frequency filter was applied. This is consistent with the expected rarity of *de novo* variants. No functional-effect filter was applied. Variants spanned a broad range of consequences, including missense, synonymous, and loss-of-function effects such as frameshift and stop-gained variants. Genetic features were assigned based on the gene in which each participant’s variant occurred.

From this resource, we extracted a set of gene-level annotations capturing distinct aspects of genetic intolerance, expression, and disease relevance. These included the probability of loss-of-function intolerance (pLI), derived from the Exome Aggregation Consortium (ExAC) (Lek et al., 2016). We also incorporated the loss-of-function observed/expected upper bound fraction (LOEUF), binned into discrete categories, derived from the Genome Aggregation Database (gnomAD) (Karczewski et al., 2020). Lower LOEUF values and higher pLI scores indicate stronger selective constraints. This suggests that loss-of-function mutations in such genes are likely deleterious (Karczewski et al., 2020).

We further incorporated the proportion expressed across transcripts (pExt), computed using the Human Brain Development Resource (HBDR) fetal brain expression dataset (Cummings et al., 2020). Higher pExt values indicate stronger fetal brain-specific expression, which may reflect greater neurodevelopmental relevance.

Additionally, we used a continuous ASD-associated risk gene score (Arisk), reflecting the degree to which a gene has been implicated in autism risk through large-scale exome sequencing studies integrating both *de novo* and inherited variant data (Zhou et al., 2020). These genetic features were mapped to each SSC participant based on their single qualifying *de novo* variant and used in combination with behavioral measures for model training and evaluation.

### 2.3 Data Preprocessing

Data preprocessing was carried out with the dual goal of preserving the most informative features while maintaining sufficient sample size for modeling. Features with more than 30% missingness were excluded, ensuring that retained predictors were consistently available across participants. After merging behavioral and genetic data, participants without genetic information were excluded, resulting in final sample sizes of 397, 466, and 1157 for Module 1, Module 2, and Module 3, respectively.

### 2.4 Machine Learning prediction of ADOS module-specific CSS

To capture the wide variability in data structure and potential predictive patterns, we trained a diverse set of 12 machine learning models encompassing linear, tree-based, and ensemble approaches. The linear models included Linear, Ridge (Hoerl & Kennard, 1970), Lasso (Tibshirani, 1996), and Elastic Net (Zou & Hastie, 2005) regression models. The tree-based algorithms included Decision Tree (Breiman et al., 1984) and Random Forest (Breiman, 2001). The ensemble methods included Gradient Boosting (Friedman, 2001), AdaBoost (Freund & Schapire, 1997), and XGBoost (Chen & Guestrin, 2016). In addition to these, we evaluated Support Vector Regression (SVR), (Drucker et al., 1997), K-Nearest Neighbors (Cover & Hart, 1967) and Multilayer Perceptron (MLP) (Rumelhart et al., 1986). The goal was not only to determine the most effective predictive model, but also to assess the stability and consistency of observed features and outcome relationships across modeling paradigms.

For each module, data were randomly divided into training and testing sets using a 70:30 split. Hyperparameters were tuned within the training portion using cross-validation, after which final models were trained and evaluated on the held-out test set. Two modeling stages were conducted. The first used only behavioral questionnaire features, while the second incorporated genetic features to assess whether their inclusion improved predictive accuracy.

### 2.5 Prediction evaluation metrics

Model performance was evaluated using two complementary metrics, Mean Absolute Error (MAE) and Pearson’s correlation coefficient (r) between the predicted and observed ADOS CSS. MAE provides an interpretable measure of the average absolute deviation between predicted and actual CSS values. It reflects the typical magnitude of prediction errors, irrespective of their direction. A lower MAE indicates closer alignment between model predictions and true values. MAE can be formulated as below in equation 1.

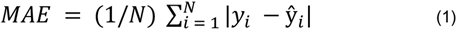

N represents the number of test samples, and *y_i_* and ŷ*_i_* represent the true and predicted severity scores respectively.

However, because CSS values are restricted to a narrow range, small absolute differences can inflate or compress the apparent performance differences between models. For instance, an MAE of 1.5 may correspond to a relatively large deviation on this scale, while still reflecting strong ordinal agreement. Thus, MAE alone may not fully capture a model’s ability to preserve meaningful patterns in severity ranking.

To address this limitation, Pearson’s r was also computed to quantify the linear association between predicted and observed scores. This metric captures how well the model preserves inter-individual variation in severity, independent of small absolute offsets. It is formulated as below in equation 2.

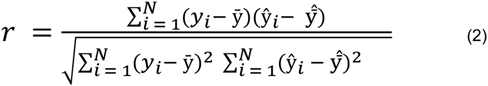

A higher correlation indicates that the model effectively retains the relative ranking of severity across individuals, even when the absolute predictions are slightly misaligned. Both metrics were reported for every model and module, under both the behavioral-only and the combined behavioral-plus-genetic feature sets, to provide a comprehensive assessment of predictive accuracy and pattern consistency.

## 3. RESULTS

### 3.1 Participant Characteristics Across ADOS Modules

Participant characteristics across ADOS Modules 1, 2, and 3 are shown in Table 1. ASD Participants in Module 3 were older than those in Modules 1 and 2 as expected. Mean ages were as follows: Module 1, 93.0 months; Module 2, 85.5 months; Module 3, 117.2 months (**Table 1**). Similarly, IQ increased significantly across modules, also consistent with the developmental expectations of the module system: Module 1 had the lowest mean IQ of 43.4, while Module 3 had the highest mean IQ of 93.2. Sex distribution also differed across modules (**Table 1**). Females accounted for 14.4% of participants in Module 1, 16.5% in Module 2, and 11.6% in Module 3. Compared with Module 3, Modules 1 and 2 included slightly higher proportions of African American, Asian, and other racial groups. (**Table 1**).

**Table 1.** Summary of participant demographics across SSC ADOS Modules 1–3. Participant characteristics are reported as mean and standard deviation for continuous variables and count percentage for categorical variables. Group differences across modules were assessed using the Kruskal–Wallis test for continuous variables and the chi-square test for categorical variables.

| **Variables** | **Module 1<br>(N=397)** | **Module 2<br>(N=466)** | **Module 3<br>(N=1157)** | **P-value** |
|---|---|---|---|---|
| Age, mean (SD) | 93.0 (41.0) | 85.5 (37.1) | 117.2 (38.8) | < 0.001<sup>1</sup> |
| IQ, mean (SD) | 43.4 (19.4) | 74.5 (20.9) | 95.2 (19.2) | < 0.001<sup>1</sup> |
| Sex |  |  |  |  |
| Male, N (%) | 340 (85.6%) | 389 (83.5%) | 1023 (88.4%) | 0.009<sup>2</sup> |
| Female, N (%) | 57 (14.4%) | 77 (16.5%) | 134 (11.6%) |  |
| Race |  |  |  |  |
| African American, N (%) | 32 (8.1%) | 26 (5.6%) | 28 (2.4%) | < 0.001<sup>2</sup> |
| Asian, N (%) | 29 (7.3%) | 27 (5.8%) | 28 (2.4%) |  |
| White, N (%) | 265 (66.8%) | 343 (73.6%) | 981 (84.8%) |  |
| Other, N (%) | 72 (18.1%) | 70 (15.2%) | 119 (10.5%) |  |

<sup>**1**</sup>Kruskal Wallis p-value; <sup>**2**</sup>Chi-Square p-value.

### 3.2 ADOS CSS Distribution by Module

CSS scores are scaled within developmental modules; consistent with this, the distributions of CSS values differed substantially across modules (**Figure 1**). Mean CSS was 7.0 (SD = 1.6) for Module 1, 7.8 (SD = 1.7) for Module 2, and 7.2 (SD = 1.8) for Module 3. A one-way ANOVA confirmed that mean CSS differed significantly across modules (p < 0.001). Based on these differing distributions, pooling all participants into a single model would risk conflating these module-specific differences and masking meaningful developmental patterns. We therefore trained models separately for Module 1, Module 2, and Module 3. By modeling each module independently, we ensure that predictions are evaluated within the appropriate developmental context and that the model learns patterns relevant to that module’s scoring scale.

**Figure 1.**
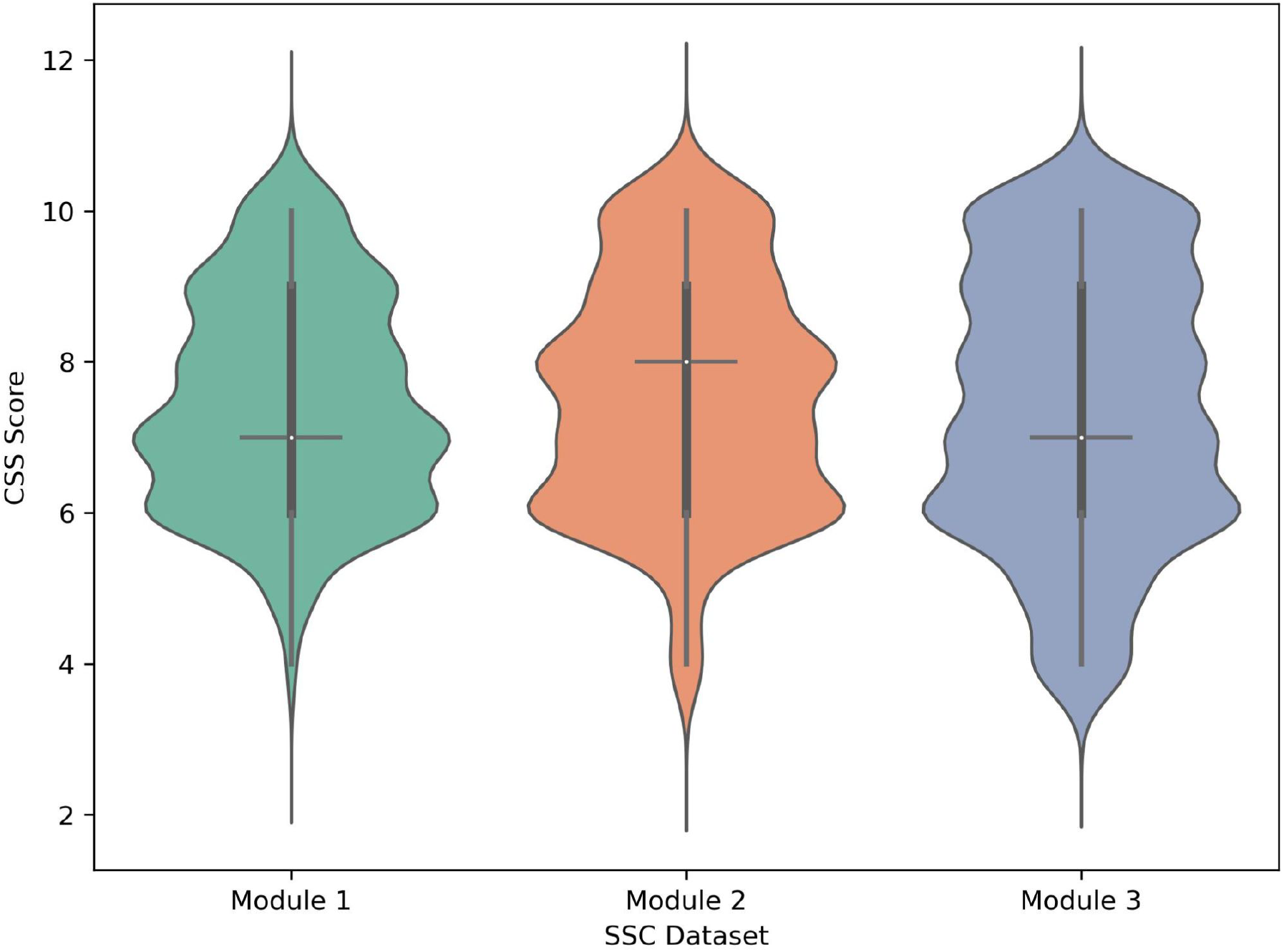
Module-wise distribution of ADOS Calibrated Severity Scores (CSS). Violin plots depict the distribution of CSS values for Modules 1–3. One-way ANOVA confirmed that mean CSS differed significantly across modules (p < 0.001)

### 3.3 ML prediction of module-specific ADOS CSS with and without genetic predictors

We evaluated 12 machine learning algorithms to predict ADOS CSS. These included Linear Regression, Ridge Regression (Hoerl and Kennard, 1970), Lasso Regression (Tibshirani, 1996), Elastic Net (Zou and Hastie, 2005), Decision Tree (Breiman et al., 1984), Random Forest (Breiman, 2001), AdaBoost (Freund and Schapire, 1997), Gradient Boosting (Friedman, 2001), XGBoost (Chen and Guestrin, 2016), Support Vector Regression (Drucker et al., 1997), K-Nearest Neighbors (Cover and Hart, 1967), and Multilayer Perceptron (MLP) (Rumelhart et al., 1986).

For each module, the dataset was randomly split into training and testing sets using a 70–30 split while maintaining the CSS distribution. In Module 1, 278 samples were used for training and 119 for testing. The mean CSS in the training set was 7.1 (SD = 1.6) and in the testing set was 7.0 (SD = 1.7). In Module 2, 326 samples were used for training and 140 for testing. The mean CSS in the training set was 7.8 (SD = 1.7) and in the testing set was 7.7 (SD = 1.8). In Module 3, 810 samples were used for training and 347 for testing. The mean CSS in the training set was 7.4 (SD = 1.8) and in the testing set was 7.3 (SD = 1.9). The distributions of CSS remained consistent between training and testing sets across all modules.

Models were trained under two feature sets. The first feature set included behavioral predictors only. Behavioral predictors included individual questions from standardized instruments such as the Social Responsiveness Scale (SRS), Repetitive Behavior Scale– Revised (RBS-R), Vineland Adaptive Behavior Scales, and related cognitive and behavioral assessments. The second feature set included genetic predictors along with behavioral predictors. This design allowed us to directly assess the incremental contribution of genetic information to prediction performance. Genetic predictors included gene-level constraint and expression-based measures such as pLI (Lek et al., 2016), LOEUF, and pExt. These capture intolerance to loss-of-function variation and brain-specific expression patterns.

Model performance was evaluated using mean absolute error (MAE) and Pearson correlation coefficient. MAE represents the average absolute difference between predicted and observed CSS values. Lower MAE values indicate greater prediction accuracy. Pearson correlation reflects concordance between predicted and observed scores.

Figures 2-4 present MAE and Pearson r for each model, comparing behavioral-only features against behavioral-plus-genetic features. In Module 1, the inclusion of genetic predictors improved performance across most models (**Figure 2**). Ensemble-based approaches showed the strongest results. Random Forest achieved the lowest MAE. MAE decreased from 1.189 without genetic predictors to 1.051 with genetic predictors added. Gradient Boosting and AdaBoost also showed reduced MAE values. Gradient Boosting decreased from 1.291 to 1.096. AdaBoost decreased from 1.204 to 1.066. These models also showed improved Pearson correlations with observed CSS scores when genetic predictors were added. Random Forest correlation increased from 0.204 to 0.340. AdaBoost correlation increased from 0.130 to 0.320. Gradient Boosting correlation increased from 0.014 to 0.300.

**Figure 2.**
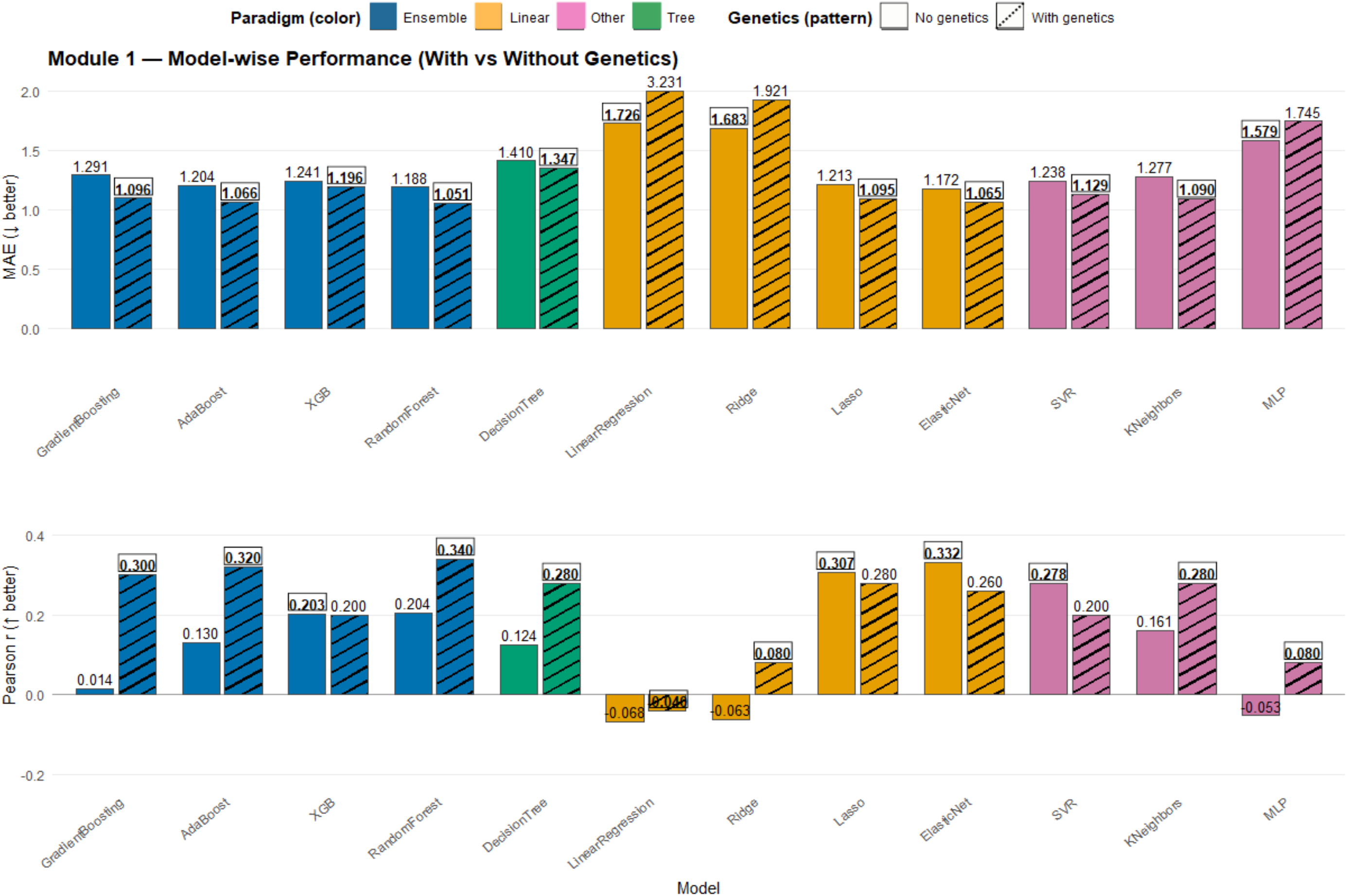
Model-wise prediction performance in Module 1 with and without genetic features. Model performance was compared across 12 machine learning algorithms using models without (solid bars) and with (hatched bars) genetic features. Colors denote model types (ensemble, linear, tree-based, and other). The upper panel shows mean absolute error (MAE; lower is better), and the lower panel shows Pearson’s correlation coefficient (r; higher is better).

In Module 2, genetic predictors continued to improve predictive performance, although the magnitude of improvement was smaller compared to Module 1 (**Figure 3**). Ensemble-based models again demonstrated the strongest overall performance. Random Forest and XGBoost achieved the highest correlations, with r values of 0.45 and 0.49, respectively. Both models also maintained relatively low MAE values. In contrast, linear approaches such as Linear Regression and Ridge Regression showed higher prediction error. These models also demonstrated limited improvement after the inclusion of genetic predictors.

**Figure 3.**
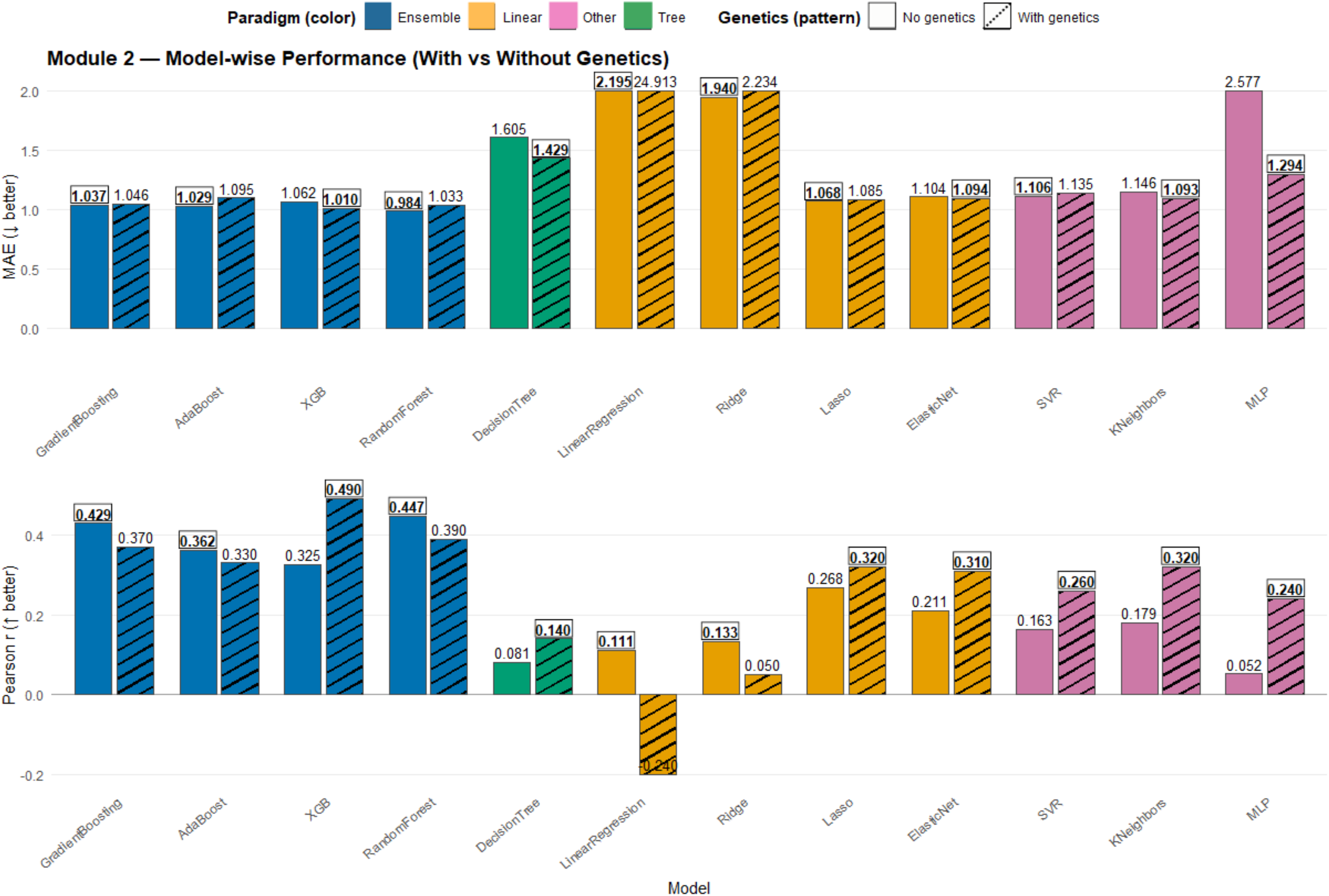
Model-wise performance in Module 2 with and without genetic features. Model performance was compared across 12 machine learning algorithms using models without (solid bars) and with (hatched bars) genetic features. Colors denote model types (ensemble, linear, tree-based, and other). The upper panel shows mean absolute error (MAE; lower is better), and the lower panel shows Pearson’s correlation coefficient (r; higher is better).

**Figure 4.**
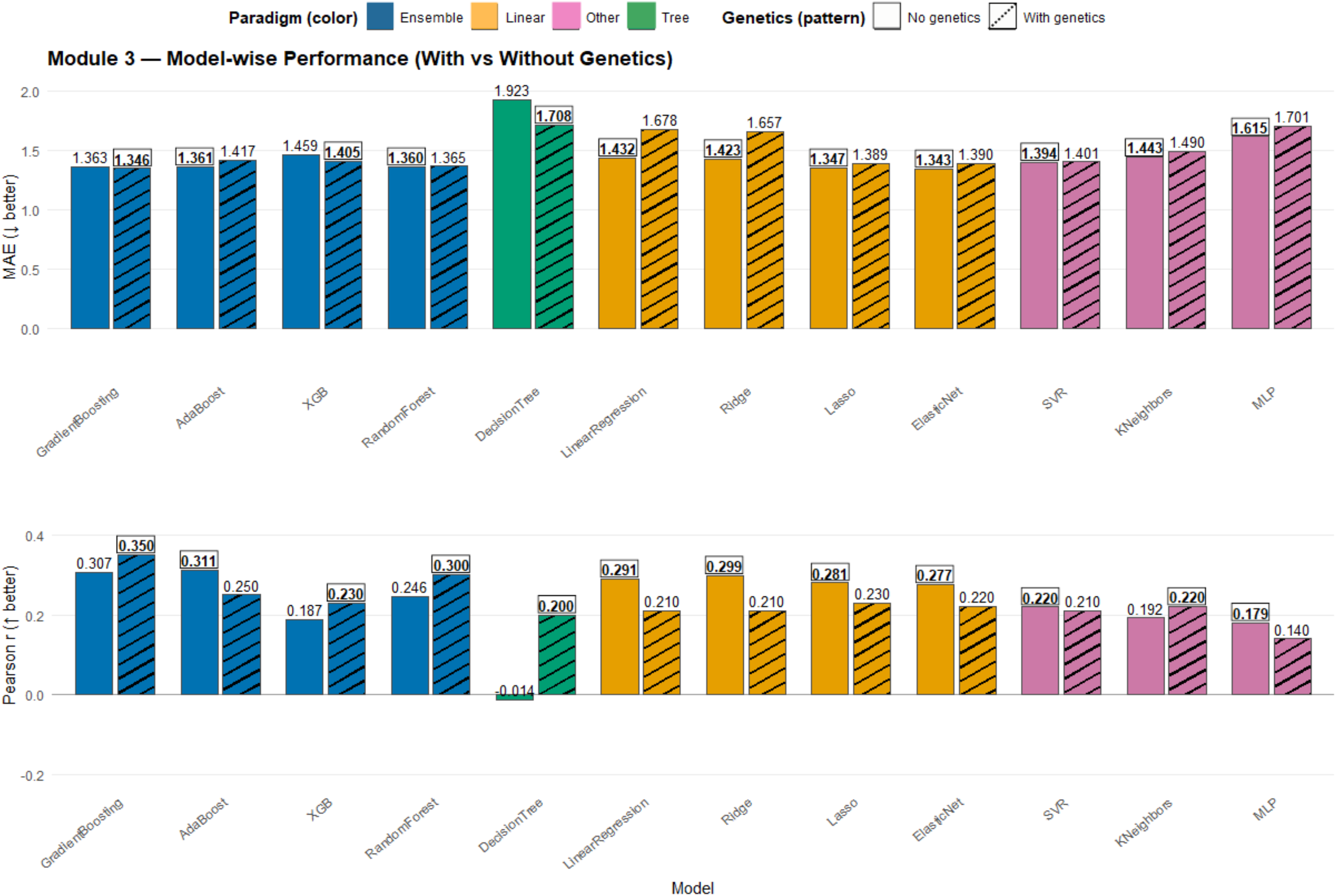
Model-wise performance in Module 3 with and without genetic features. Model performance was compared across 12 machine learning algorithms using models without (solid bars) and with (hatched bars) genetic features. Colors denote model types (ensemble, linear, tree-based, and other). The upper panel shows mean absolute error (MAE; lower is better), and the lower panel shows Pearson’s correlation coefficient (r; higher is better).

In Module 3, the contribution of genetic predictors was comparatively modest (**Figure 4**). Ensemble-based models continued to outperform linear and kernel-based approaches. Improvements associated with genetic information were less pronounced than in earlier developmental modules. Pearson correlations among the top-performing models ranged from approximately 0.20 to 0.35. This indicated moderate predictive performance overall. This modest contribution of genetic predictors likely reflects the well-documented relationship between genetic architecture and clinical presentation. Higher-functioning individuals, who make up much of Module 3, are known to carry fewer high-impact *de novo* variants than more severely affected individuals (Robinson et al., 2014). There may therefore have been less genetic signal available to detect in this group, rather than a true decline in the relevance of genetic factors at this developmental stage. Behavioral features accounted for a larger proportion of predictive signal in Module 3. Genetic predictors provided only smaller incremental improvements.

In summary, genetic predictors improved the prediction of CSS, particularly in Module 1 and 2, and provided complementary information to behavioral measures. Their contribution was most pronounced in Module 1 and diminished in later modules. Across all modules, ensemble-based approaches demonstrated the most robust performance. Random Forest, Gradient Boosting, and AdaBoost consistently outperformed linear and kernel-based models.

### 3.4 Measures with predictive influence of ADOS CSS scores

Beyond predictive accuracy, another goal of this study was to identify which genetic and behavioral features most strongly influence ADOS CSS predictions across the best-performing models in Modules 1, 2, and 3. SHAP (SHapley Additive exPlanations) provides a unified framework to quantify the contribution of each feature to individual model predictions. SHAP analyses of our top-performing models, Random Forest (Module 1 and 3) and XGBoost (Module 2), provided clear, structured patterns in feature importance.

Across both models, genetic predictors, particularly pLI (gene intolerance), emerged as key contributors to model predictions. Higher pLI values indicate greater intolerance to loss-of-function variants and were associated with increased predicted severity. We also incorporated the proportion expression across tissues (pExt), a transcript-level metric reflecting tissue-specific gene expression. Higher pExt values in brain tissues suggest greater neurodevelopmental relevance (Cummings et al., 2020).

Among behavioral predictors of CSS in Module 1 using Random Forest (**Figure 5**), Vineland-II Written V-score, nonverbal IQ, and Vineland-II Written Age Equivalent had the highest feature importance rankings. It is noted that Vineland-II Written scores are at the floor in Module 1 due to participants’ pre-verbal status. These features showed positive SHAP values for lower scores, indicating that reduced adaptive and cognitive abilities increase predicted CSS, corresponding to greater symptom severity. Measures reflecting impulsivity and attentional control (e.g., Impulsivity Score and Ignores Instructions question from ABC) also contributed meaningfully.

**Figure 5.**
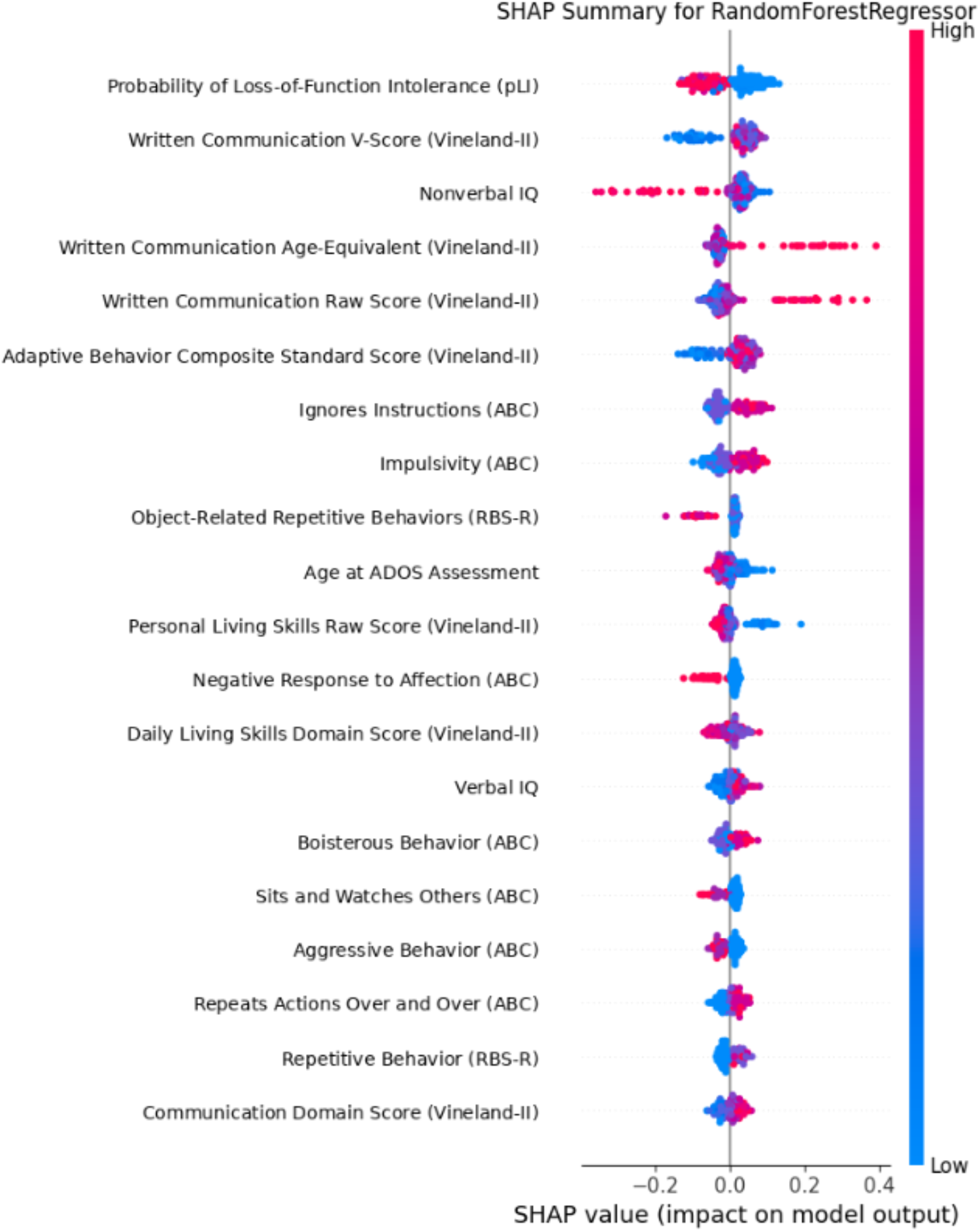
SHAP Summary Plot for Random Forest Model on predicting ADOS CSS in Module 1. SHAP summary plot illustrating the contribution of adaptive functioning, cognitive, and genetic features to predicted ADOS Calibrated Severity Scores in Module 1. Each dot represents an individual observation, colored by feature value. Positive SHAP values indicate increased predicted symptom severity.

The XGBoost model (**Figure 6**) in Module 2 exhibited a slightly different feature ranking but a complementary pattern. Genetic predictors such as pLI again appeared among the top features, indicating consistent importance across model types. Cognitive measures such as verbal IQ, full-scale IQ, and Vineland-II receptive language measures were also prominent. Social-behavioral variables from the SRS, including difficulty making friends and response patterns in communication, contributed to predictions.

**Figure 6.**
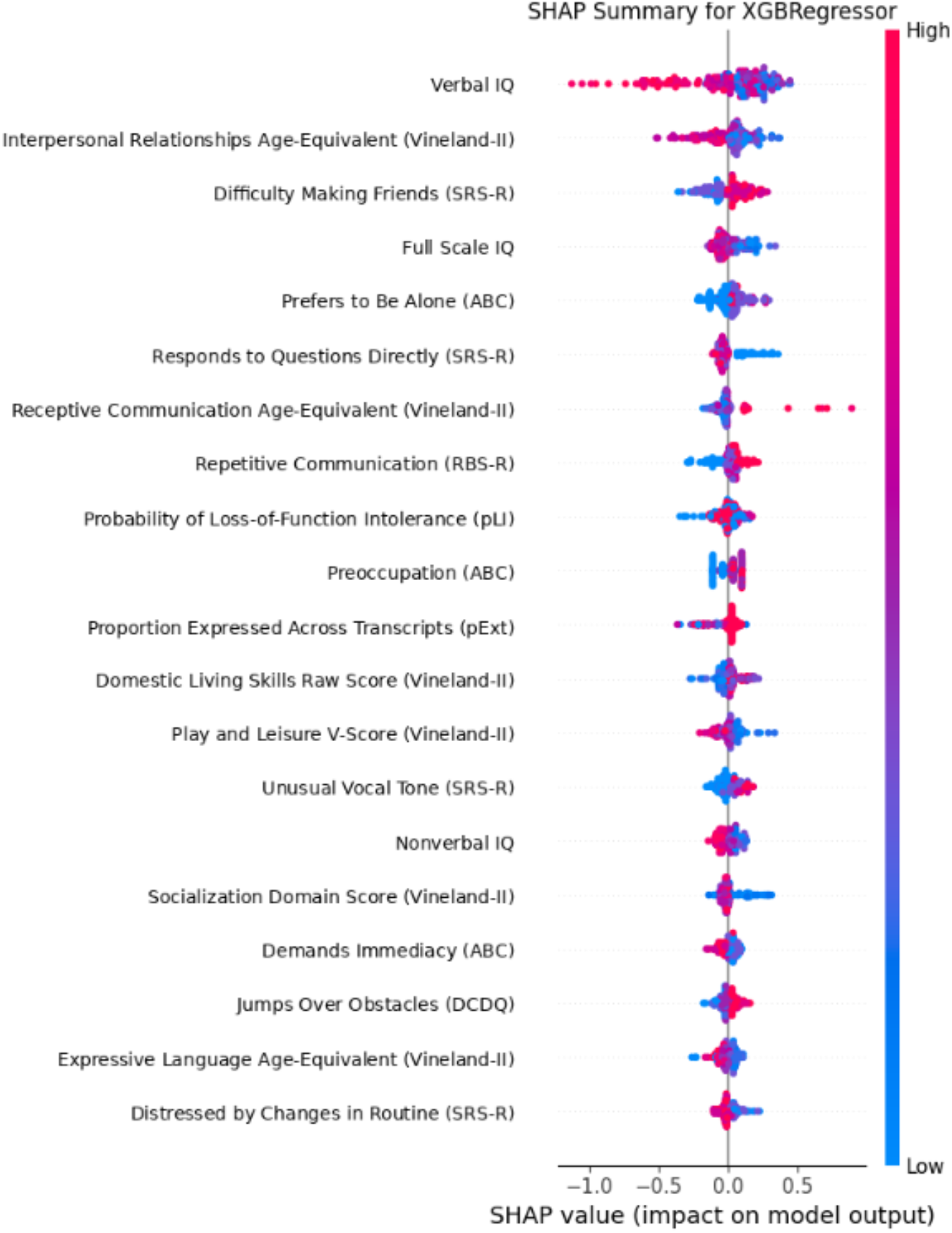
SHAP Summary Plot for XG Boost Model on predicting ADOS CSS in Module 2. SHAP summary plot illustrating the contribution of adaptive functioning, cognitive, and genetic features to predicted ADOS Calibrated Severity Scores in Module 2. Each dot represents an individual observation, colored by feature value. Positive SHAP values indicate increased predicted symptom severity.

In Module 3, we performed SHAP analysis on the Random Forest model (**Figure 7**). The predictive structure shifted more strongly toward higher-order behavioral and social-cognitive features. Unlike Module 1, where genetic intolerance measures such as pLI showed strong predictive influence, Module 3 predictions were dominated primarily by verbal IQ, adaptive social functioning, and nuanced social communication behaviors. SHAP analysis demonstrated that verbal IQ was the most influential contributor in Module 3, with lower verbal ability consistently associated with increased predicted CSS.

**Figure 7.**
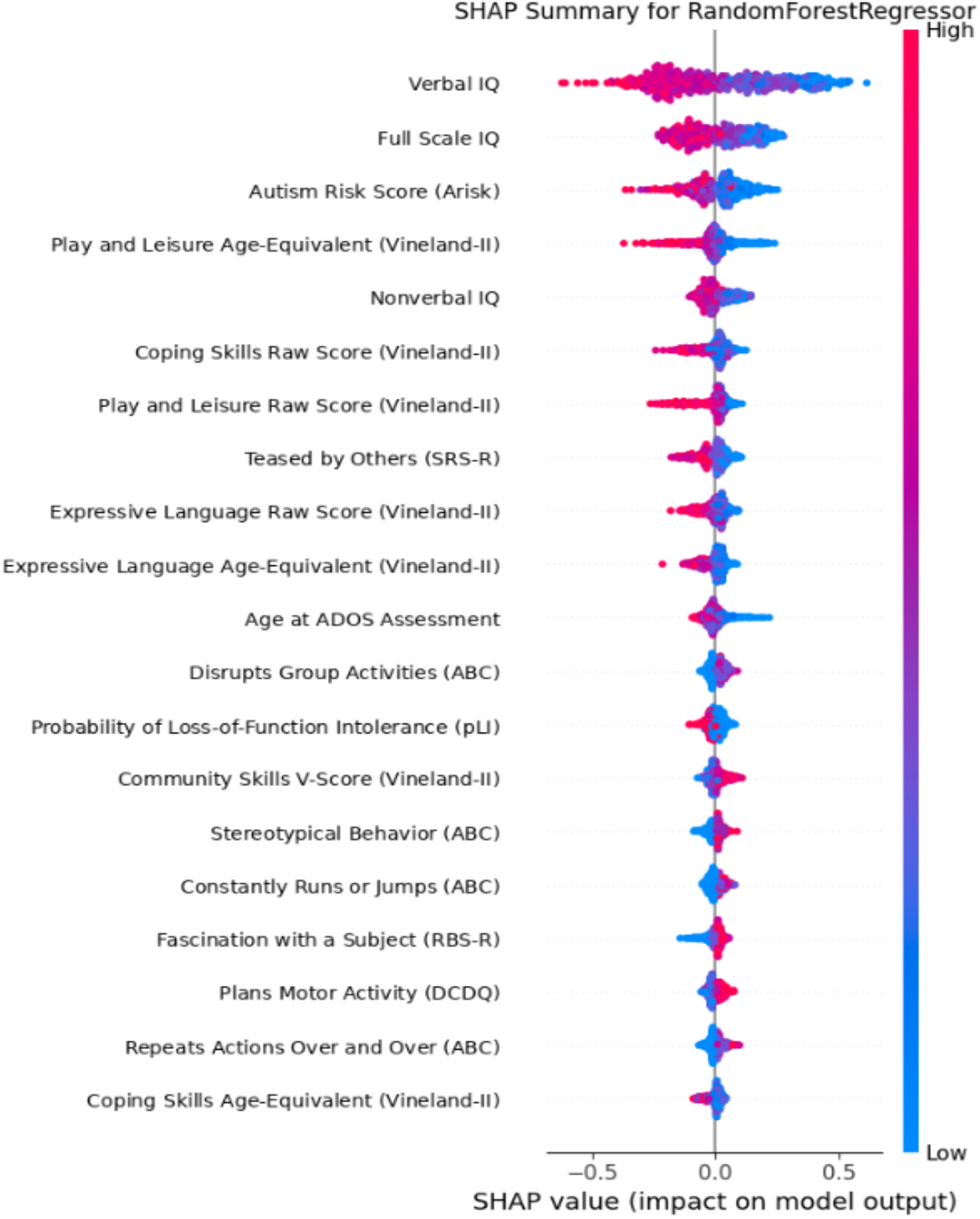
SHAP Summary Plot for Random Forest Model on predicting ADOS CSS in Module 3. SHAP summary plot illustrating the contribution of adaptive functioning, cognitive, and genetic features to predicted ADOS Calibrated Severity Scores in Module 3. Each dot represents an individual observation, colored by feature value. Positive SHAP values indicate increased predicted symptom severity.

SHAP values indicate both feature importance and direction of influence. Genetic predictors such as pLI and pExt shifted predictions upward, with higher values associated with increased predicted CSS. Lower Vineland scores increased predicted CSS, while higher IQ values were associated with lower predicted severity.

## 4. DISCUSSION

In this study, we developed and evaluated machine learning models to predict quantitative severity of autism characteristics as measured by the ADOS CSS. Both common behavioral questionnaire measures and gene-level annotations were used. Across developmental modules, ensemble-based machine learning models, notably Random Forest, Gradient Boosting, AdaBoost, and XGBoost, showed the best predictive performance. Adding genetics to the predictive models increased our ability to identify children with a higher likelihood of increased autism symptom severity, particularly among those with little or no expressive language. The incremental value of genetic predictors was most evident in Module 1, which also happens to correspond to the youngest age group in the SSC on average. Module 1 includes children with little or no phrase speech. This may explain why the inclusion of genetic features resulted in the largest model performance gains in Module 1.

The heritability of ASD has been repeatedly demonstrated to be high, with estimates ranging from 64% to 91% (Tick et al., 2016). Prior work has shown that the genetic architecture of ASD varies systematically with clinical presentation. This aligns with our findings that genetic predictors improve CSS prediction most substantially in early developmental stages, where symptom severity may be more tightly coupled with underlying biological mechanisms, reflected in gene-level intolerance scores such as pLI and LOEUF. This interpretation is supported by previous work demonstrating that greater genetic burden, particularly intolerance to loss-of-function variation, is associated with more severe neurodevelopmental phenotypes, including ASD (Robinson et al., 2014; Lek et al., 2016). SHAP analysis improves interpretability and demonstrates that the models do not function as black boxes. This also highlights how specific genetic and behavioral features contribute to predictions. Our SHAP analyses similarly identified pLI as one of the most influential predictors across the best-performing models, suggesting that gene intolerance contributes meaningfully to variation in autism symptom severity.

Although this study focused on behavioral and genetic predictors, existing literature makes it clear that environmental factors also play an important role in ASD risk and severity. Prenatal exposures, maternal health, perinatal complications, and early-life environments have all been implicated in shaping phenotypic variability (Modabbernia et al., 2017). Importantly, gene-environment interactions (G×E) are increasingly recognized as critical in determining symptom expression and developmental outcomes (Tordjman et al., 2014; Persico & Napolioni, 2022). Future research should incorporate environmental exposures alongside genetic and behavioral measures to determine whether modeling gene–environment interactions further improves prediction of autism symptom severity across developmental stages.

Importantly, SHAP-based feature importance revealed module-specific patterns that align closely with the developmental design of the ADOS assessment. In Module 1, nonverbal IQ emerged as a top predictor of CSS score. This is consistent with the structure of Module 1, which is designed for children with little or no phrase speech. In contrast, in Module 2, verbal IQ was among the most influential predictors. Module 2 includes children who have phrase speech but are not yet verbally fluent. The predominance of verbal IQ in Module 3 is also developmentally consistent with the ADOS framework. Module 3 is specifically designed for verbally fluent individuals, in whom symptom severity is assessed through increasingly complex social communication and reciprocal interaction rather than early language acquisition. Consequently, verbal ability becomes a stronger determinant of variation in CSS than in earlier modules.

The behavioral features driving Module 3 predictions differed qualitatively from those observed in earlier modules. Rather than broad developmental impairments, the model relied more heavily on subtle impairments in social participation and pragmatic communication. This suggests that, in older and more verbally fluent individuals, autism severity is increasingly reflected through complex social-cognitive and interpersonal functioning rather than foundational developmental deficits alone.

These findings provide strong empirical support for modeling ADOS CSS separately by module. The predictors driving model performance are not uniform across developmental stages. Instead, they reflect the underlying clinical and developmental distinctions encoded within the ADOS framework. Pooling all participants into a single model would obscure these meaningful differences and reduce both predictive accuracy and interpretability.

Our finding that genetic features contributed less to predictive performance in Modules 2 and 3 may reflect this developmental shift. As children grow older, accumulated environmental and experiential factors increasingly shape observed symptom severity, while the relative influence of genetic liability on phenotype may become less pronounced. In particular, children who progress to the more advanced ADOS modules have often experienced years of intensive early intervention, educational support, speech and language therapy, and other developmental interventions. These cumulative experiences can substantially influence language, social communication, and adaptive functioning, potentially attenuating the direct relationship between underlying genetic predisposition and observed clinical severity. Thus, in later developmental stages, behavioral presentation may increasingly reflect the combined effects of genetic susceptibility and accumulated environmental and therapeutic influences.

## Data Availability

Approved researchers can obtain the SSC dataset described in this study by applying at https://base.sfari.org.

## 3. ACKNOWLEDGMENTS

This work was supported by National Institute of Mental Health (R01MH143902). We are grateful to all the families in SSC, the SSC clinical sites and SSC staff. We appreciate obtaining access to the SSC phenotypic data set on SFARI Base. Approved researchers can obtain the SSC dataset described in this study by applying at https://base.sfari.org.

## 5. ETHICS STATEMENT

The secondary analysis of de-identified data in SSC was approved by the Institutional Review Board of the University of Southern California (IRB Protocol No. HS-26-00184).

## REFERENCES

Lord, C., Rutter, M., DiLavore, P. C., Risi, S., Gotham, K., & Bishop, S. (2012). Autism Diagnostic Observation Schedule, Second Edition (ADOS-2). Western Psychological Services.

Gotham, K., Pickles, A., & Lord, C. (2009). Standardizing ADOS scores for a measure of severity in autism spectrum disorders. Journal of Autism and Developmental Disorders, 39(5), 693–705. 10.1007/s10803-008-0674-3

Constantino, J. N., & Gruber, C. P. (2005). Social Responsiveness Scale (SRS) manual. Western Psychological Services.

Bodfish, J. W., Symons, F. J., Parker, D. E., & Lewis, M. H. (2000). Varieties of repetitive behavior in autism: Comparisons to mental retardation. Journal of Autism and Developmental Disorders, 30(3), 237–243. 10.1023/A:1005596502855

Tick, B., Bolton, P., Happé, F., Rutter, M., & Rijsdijk, F. (2016). Heritability of autism spectrum disorders: A meta-analysis of twin studies. Journal of Child Psychology and Psychiatry, 57(5), 585–595. 10.1111/jcpp.12499

Grove, J., Ripke, S., Als, T. D., Mattheisen, M., Walters, R. K., Won, H., et al. (2019). Identification of common genetic risk variants for autism spectrum disorder. Nature Genetics, 51(3), 431–444. 10.1038/s41588-019-0344-8

Trost, B., Thiruvahindrapuram, B., Chan, A. J. S., Engchuan, W., Higginbotham, E. J., Howe, J. L., et al. (2022). Genomic architecture of autism from comprehensive whole-genome sequence annotation. Cell, 185(23), 4409–4427.e18. 10.1016/j.cell.2022.10.009

Zhou, X., Feliciano, P., Shu, C., Wang, T., Astrovskaya, I., Hall, J. B., et al. (2022). Integrating de novo and inherited variants in 42,607 autism cases identifies mutations in new moderate-risk genes. Nature Genetics, 54(9), 1305–1319. 10.1038/s41588-022-01148-2

Duda, M., Haber, N., Daniels, J., & Wall, D. P. (2017). Crowdsourced validation of a machine-learning classification system for autism and ADHD. Translational Psychiatry, 7(5), e1133. 10.1038/tp.2017.86

Farooq, M. S., Tehseen, R., Sabir, M., et al. (2023). Detection of autism spectrum disorder (ASD) in children and adults using machine learning. Scientific Reports, 13, 9605. 10.1038/s41598-023-35910-1

Rajagopalan, S. S., Zhang, Y., Yahia, A., & Tammimies, K. (2024). Machine learning prediction of autism spectrum disorder from a minimal set of medical and background information. JAMA Network Open, 7(8), e2429229. 10.1001/jamanetworkopen.2024.29229

Briguglio, M., Turriziani, L., Currò, A., Gagliano, A., Di Rosa, G., Caccamo, D., Tonacci, A., & Gangemi, S. (2023). A machine learning approach to the diagnosis of autism spectrum disorder and multi-systemic developmental disorder based on retrospective data and ADOS-2 score. Brain Sciences, 13(6), 883. 10.3390/brainsci13060883

Fischbach, G. D., & Lord, C. (2010). The Simons Simplex Collection: A resource for identification of autism genetic risk factors. Neuron, 68(2), 192–195. 10.1016/j.neuron.2010.10.006

Sparrow, S. S., Cicchetti, D. V., & Balla, D. A. (2005). Vineland Adaptive Behavior Scales, Second Edition (Vineland-II). Pearson Assessments.

Aman, M. G., & Singh, N. N. (1985). Aberrant Behavior Checklist manual. Slosson Educational Publications. (placeholder — please confirm this is the edition you intend to cite)

Shumway, S., Farmer, C., Thurm, A., Joseph, L., Black, D., & Golden, C. (2012). The ADOS calibrated severity score: Relationship to phenotypic variables and stability over time. Autism Research, 5(4), 267–276. 10.1002/aur.1238

Lek, M., Karczewski, K. J., Minikel, E. V., Samocha, K. E., Banks, E., Fennell, T., et al. (2016). Analysis of protein-coding genetic variation in 60,706 humans. Nature, 536(7616), 285–291. 10.1038/nature19057

Karczewski, K. J., Francioli, L. C., Tiao, G., Cummings, B. B., Alföldi, J., Wang, Q., et al. (2020). The mutational constraint spectrum quantified from variation in 141,456 humans. Nature, 581(7809), 434–443. 10.1038/s41586-020-2308-7

Cummings, B. B., et al. (2020). [Human Brain Development Resource fetal brain expression dataset]. I could not independently verify the exact title/journal for this citation — please double-check this one in your source material.

Zhou, X., et al. (2020). [ASD-associated risk gene score]. This may be the same underlying study as #8 above (Zhou et al., 2022), which may have circulated as a 2020 preprint — please confirm against your reference manager.

Hoerl, A. E., & Kennard, R. W. (1970). Ridge regression: Biased estimation for nonorthogonal problems. Technometrics, 12(1), 55–67. 10.1080/00401706.1970.10488634

Tibshirani, R. (1996). Regression shrinkage and selection via the lasso. Journal of the Royal Statistical Society: Series B, 58(1), 267– 288.

Zou, H., & Hastie, T. (2005). Regularization and variable selection via the elastic net. Journal of the Royal Statistical Society: Series B, 67(2), 301–320. 10.1111/j.1467-9868.2005.00503.x

Breiman, L., Friedman, J. H., Olshen, R. A., & Stone, C. J. (1984). Classification and regression trees. Wadsworth.

Breiman, L. (2001). Random forests. Machine Learning, 45(1), 5–32. 10.1023/A:1010933404324

Friedman, J. H. (2001). Greedy function approximation: A gradient boosting machine. The Annals of Statistics, 29(5), 1189–1232. 10.1214/aos/1013203451

Freund, Y., & Schapire, R. E. (1997). A decision-theoretic generalization of on-line learning and an application to boosting. Journal of Computer and System Sciences, 55(1), 119–139. 10.1006/jcss.1997.1504

Chen, T., & Guestrin, C. (2016). XGBoost: A scalable tree boosting system. In Proceedings of the 22nd ACM SIGKDD International Conference on Knowledge Discovery and Data Mining (pp. 785–794). 10.1145/2939672.2939785

Drucker, H., Burges, C. J. C., Kaufman, L., Smola, A., & Vapnik, V. (1997). Support vector regression machines. In Advances in Neural Information Processing Systems (Vol. 9, pp. 155–161). MIT Press.

Cover, T., & Hart, P. (1967). Nearest neighbor pattern classification. IEEE Transactions on Information Theory, 13(1), 21–27. 10.1109/TIT.1967.1053964

Rumelhart, D. E., Hinton, G. E., & Williams, R. J. (1986). Learning representations by back-propagating errors. Nature, 323(6088), 533–536. 10.1038/323533a0

Robinson, E. B., Samocha, K. E., Kosmicki, J. A., et al. (2014). Autism spectrum disorder severity reflects the average contribution of de novo and familial influences. Proceedings of the National Academy of Sciences, 111(42), 15161–15165. 10.1073/pnas.1409204111

Modabbernia, A., Velthorst, E., & Reichenberg, A. (2017). Environmental risk factors for autism: An evidence-based review of systematic reviews and meta-analyses. Molecular Autism, 8, 13. 10.1186/s13229-017-0121-4

Tordjman, S., Somogyi, E., Coulon, N., Kermarrec, S., Cohen, D., Bronsard, G., et al. (2014). Gene × environment interactions in autism spectrum disorders: Role of epigenetic mechanisms. Frontiers in Psychiatry, 5, 53. 10.3389/fpsyt.2014.00053

Persico, A. M., & Napolioni, V. (2013). Autism genetics. Behavioural Brain Research, 251, 95–112. 10.1016/j.bbr.2013.06.012

